# Development and Validation of an Automated Image-Based Method for Prognostic Stratification in Lung Adenocarcinoma using Immunohistochemical Markers

**DOI:** 10.64898/2026.10.22.26366204

**Authors:** Filippo Carraro, María Sangüesa, Amaia Irisarri, Daniel Orive, Cristina Sainz, Josu Salinas, Ignacio Arganda-Carreras, Alfonso Calvo, Luis M. Montuenga, Carlos Ortiz de Solórzano, Mikel Ariz

## Abstract

We present an automated image-based framework for quantifying biomarker expression in immunohistochemical (IHC) images and introduce the Marker Expression Level (MEL) score as an objective alternative to visual H-scoring. MEL addresses key limitations of existing automated methods by integrating tissue-region segmentation with tumour-area–weighted marker quantification, normalization to mitigate inter-sample and inter-batch staining variability, and robust quantification of nuclear, cytoplasmic, and membrane-associated biomarkers. The complete pipeline was developed as an open-source Fiji/ImageJ tool and is publicly available.

We evaluated MEL using IHC images from 128 lung adenocarcinoma patients stained for BRCA1, GLUT1, and QKI, three markers forming a prognostic signature previously described by our group.

Tissue regions were segmented using a machine learning-based Fiji plugin, and MEL scores were calculated within tumour regions. A normalization strategy accounted for inter-TMA staining variability. Agreement with visual H-scores was assessed using Spearman correlation. Prognostic performance was evaluated using Cox regression to derive a Combined Prognostic Index (CPI) incorporating MEL scores and tumour stage.

Segmentation achieved high accuracy across tissue regions (Dice coefficients 0.69–0.99), with lower performance in highly immune-infiltrated stromal areas. MEL scores significantly correlated with visual H-scores for all markers (ρ = 0.61–0.69, p < 0.001). Normalization improved agreement for GLUT1 (ρ = 0.39 to 0.62). The derived CPI stratified patients into distinct risk groups with significant differences in disease-free and overall survival (p < 0.05), comparable to visual scoring.

MEL provides a reproducible and scalable alternative to expert visual H-scoring and standard semi- automated approaches such as QuPath. Its integration with prognostic modelling reproduced a validated survival signature with performance comparable to expert scoring, supporting clinical and translational applicability. MEL is not a marginal refinement of existing digital scoring methods, but rather a clinically oriented and biologically adaptable framework designed to improve the robustness and reproducibility of automated IHC quantification.

## INTRODUCTION

Lung cancer remains a leading cause of cancer-related mortality worldwide, with lung adenocarcinoma representing its most prevalent subtype. Despite therapeutic advances, outcomes remain poor, largely due to late-stage diagnosis, marked biological heterogeneity and resistance to novel targeted or immune-based therapies [1]. Current practical prognostic assessment relies primarily on tumour-node-metastasis (TNM) staging and histological classification, as well as other additional traits such as age, comorbidities or performance status. However, these parameters incompletely capture tumour biology and variability in clinical outcomes. The practical experience shows that within the same stage there are cases with different risk of relapse, survival or potential response to adjuvant treatment [2].

Protein expression, assessed by immunohistochemistry (IHC), represents a clinically feasible and cost-effective source of prognostic information, as it can be applied to routinely processed formalin- fixed paraffin-embedded tissue biopsies. Several protein biomarkers have been proposed, including proliferation markers (e.g., Ki-67), tumour suppressors (e.g., p65, p63), differentiation markers (e.g., TTF1), and immune-related proteins (e.g., CD8/CD3, etc.), although their real-life clinical adoption has so far been limited by variability in study design and lack of robust validation [3], [4], [5].

In previous work, our group developed an IHC prognostic signature for early-stage lung adenocarcinoma, demonstrating a significant improvement in risk stratification over TNM staging alone and offering potential clinically actionable guidance for decisions regarding adjuvant therapy [6]. The IHC staining of the three signature proteins was present either in the nucleus, the cytoplasm or the membrane. This signature was validated using well-characterized clinical cohorts, successfully addressing common limitations such as heterogeneous study design and insufficient validation. However, despite its promising prognostic performance, its implementation still relied on visual H- score assessment, which remains time-consuming and subject to inter- and intra-observer variability [7], limiting its potential clinical use. Indeed, the common practice in diagnostic Pathology still relies on visual IHC scoring of the overall staining, the percentage of positive cells (e.g. PD-L1 quantification [8]), the intensity staining levels (e.g. HER2 [9]) or a combination of the above using semi-quantitative scores, such as the H-score (e.g. ER [10]), which combines staining intensity and the percentage of positively stained cells.

To address the limitations of visual IHC scoring, several automated and computer-assisted methods have been proposed. These methods can be broadly categorized into approaches that directly quantify staining intensity at the pixel level and approaches that rely on cell or tissue segmentation prior to quantification. One representative pixel-level strategy is the pixel-wise H-score (pix H-score) proposed by Ram et al. [11], which evaluates IHC staining at the single-pixel level without requiring cell segmentation. In this approach, pixels are classified according to staining intensity, and the final score is computed digitally from the global distribution of intensities across the image, providing a more objective and reproducible alternative to manual H-scoring. Among the second group, recent Deep Learning-based frameworks, such as the convolutional neural network (CNN) approach proposed by Wen et al. [12], perform tumour region segmentation followed by automated classification of nuclei according to staining intensity categories (negative, weak, moderate, and strong). A final H-score is then computed by integrating staining intensity with the proportion of positive tumour cells, thereby reproducing the conventional semi-quantitative visual scoring process in a standardized manner. Similarly, open-source digital pathology platforms such as QuPath [13] enable customized segmentation-based quantification workflows. For example, Naso et al. [14] used QuPath cell detection algorithms to identify tumour cells and classify membrane staining intensity for automated PD-L1 scoring in non-small cell lung carcinoma. Several additional segmentation methodologies for lung histopathology images have also been proposed in recent years, including machine learning and deep learning approaches specifically designed for tumour tissue segmentation [15], [16], [17], [18].

Despite these advances, important challenges remain unresolved, including limited adaptability across different cellular compartments, sensitivity to inter-batch staining variability, and difficulties in obtaining robust and reproducible quantification of membrane-associated biomarkers. Motivated by these limitations we have designed an automated, image-based pipeline that implements the Marker Expression Level (MEL) score, a new metric that can quantify marker expression in tumours. We present this metric, compare it to the visual H-score and replicate a previously established protein- based prognostic signature [6]. By leveraging the MEL Score, we aim to enhance the accuracy and efficiency of biomarker quantification, ultimately providing a robust tool for clinical decision- making. Unlike previously proposed automated IHC quantification strategies, including pixel-wise

H-score methods, deep learning-based H-score estimation, and generic QuPath pipelines [15], [16], [17], [18], the MEL score was designed to address several unsolved limitations that hinder clinical reproducibility and retrospective translational studies. First, we introduce a machine learning-based method for tissue segmentation on a superpixel basis [19], differentiating background, tumour, stroma and immune infiltrate areas. Second, MEL integrates tumour-area–weighted IHC quantification across tissue samples, reducing sampling bias and providing a more biologically representative assessment of marker expression. Third, the method incorporates an explicit normalization strategy to compensate for inter-sample and inter-batch-staining variability, a major but often overlooked source of technical bias in long-term retrospective cohorts. Fourth, MEL is not restricted to a single cellular compartment or staining pattern, but supports robust quantification of nuclear, cytoplasmic and, relevantly, membrane-associated biomarkers such as GLUT1, whose accurate segmentation remains technically challenging in digital pathology.

## MATERIALS AND METHODS

### Patient Cohort and Inclusion Criteria

A retrospective cohort of 128 patients with surgically resected lung adenocarcinoma from Clínica Universidad de Navarra (CUN, Pamplona, Spain) was considered. Patients had stage I–III non-small cell lung cancer, underwent surgical resection at CUN, and had no previous cancer within 5 years before surgery, except non-melanoma skin cancer. Patients signed the informed consent and the project was approved by the University of Navarra Ethics Committee (Ref. 092.2012 mod 1). The cohort and the prognostic signature based on BRCA1, GLUT1, and QKI expression have been previously described and validated [6], with the corresponding prognostic index developed using multivariable Cox regression following TRIPOD guidelines [20]. For the present study, patients were required to have at least one evaluable TMA core image for each biomarker and available visual H- scores for all three markers, resulting in a final analytical cohort of 96 patients. Clinical characteristics of the final cohort are summarized in Table 1. Detailed cohort characteristics, TMA construction, immunohistochemistry, and visual scoring procedures are provided in the Supplementary Methods and in [6].

**Table 1:** Summary of clinical traits of the patient cohort used in this study. Data are presented as N (%) unless otherwise indicated. Age is presented as median (interquartile range). Percentages were calculated using the total study population (N = 89). NA: not available.

| Variable | Subgroup | N (%) |
|---|---|---|
| Age | - | 64 (57-70) |
| Gender | Male | 66 (74.2%) |
|  | Female | 23 (25.8%) |
| Smoking status | Current smoker | 62 (69.7%) |
|  | Former smoker | 13 (14.6%) |
|  | Never smoker | 14 (15.7%) |
| Pathological stage | IA | 37 (41.6%) |
|  | IB | 16 (18.0%) |
|  | IIA | 18 (20.2%) |
|  | IIB | 8 (9.0%) |
|  | IIIA | 10 (11.2%) |
| Adjuvant therapy | No | 72 (80.9%) |
|  | Yes | 7 (7.9%) |
|  | NA | 10 (11.2%) |
| Follow-up status | DF | 61 (68.5%) |
|  | PD | 26 (29.2%) |
|  | NA | 2 (2.2%) |
| Overall survival status | Alive | 64 (71.9%) |
|  | Died | 24 (27.0%) |
|  | NA | 1 (1.1%) |

### Image Preprocessing

Before tissue segmentation, background noise and illumination inhomogeneities were reduced by normalizing each image to its modal histogram intensity. The modal value was shifted to 255 by adding a constant offset to all pixels. Histogram equalization was then applied to enhance tissue contrast and facilitate segmentation. This operation was used exclusively for tissue segmentation and not for marker quantification, as it could alter the measured protein expression levels.

### Tissue Segmentation

Tissue regions were segmented using Trainable Superpixel Segmentation (TSS; Figure 1), an in- house machine learning-based plugin developed in Fiji [19], [21]. It is based on the Weka Trainable Segmentation plugin [22]. TSS classifies SLIC-generated superpixels according to their image features, reducing computational cost and sensitivity to local pixel-level noise [23].

**Figure 1:**
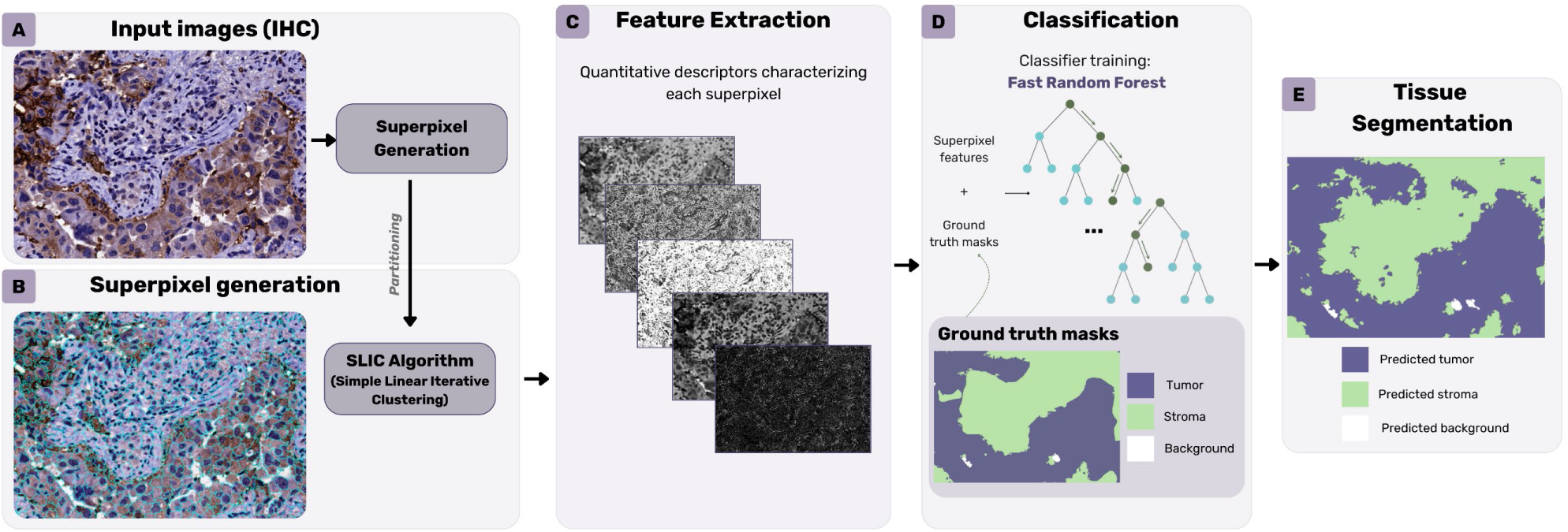
Trainable Superpixel Segmentation Pipeline.

The model was trained using pre-processed images, corresponding superpixel label maps, and ground-truth masks obtained by consensus annotation of 20 images per marker by three expert histopathology readers (D.O., C.S., and A.C.). Four tissue regions were annotated: background, tumour, stroma, and immune infiltration. Segmentation performance was evaluated by leave-one-out cross-validation using the 60 annotated images, with one image excluded for testing in each fold. Finally, a model trained on all annotated images was applied to the remaining dataset, extracting the four tissue regions for downstream image analysis.

The TSS plugin is available at https://github.com/CVPD/Trainable_Superpixel_Segmentation.

### Quantification of Marker Expression

The MEL score was calculated as the tumour-area-weighted mean DAB staining intensity across the available tumour cores of each patient:

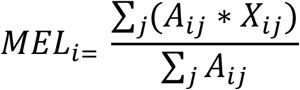

where X_i,j_ is mean staining intensity in core *j* (with *j* being 1, 2 o 3) of patient *i,* and A_i,j_ is the corresponding tumour area.

The quantification pipelines for nuclear BRCA1, membrane GLUT1, and nuclear and cytoplasmic QKI are summarized in Figure 2. For BRCA1 and nuclear QKI, colour deconvolution was used to separate the haematoxylin and DAB channels [24]. Nuclei were segmented from the haematoxylin channel by intensity thresholding and morphological post-processing. The tumour segmentation mask was then applied to retain only tumour nuclei, and mean DAB intensity was measured within the resulting mask. The patient-level MEL score was calculated using the available tumour cores.

**Figure 2:**
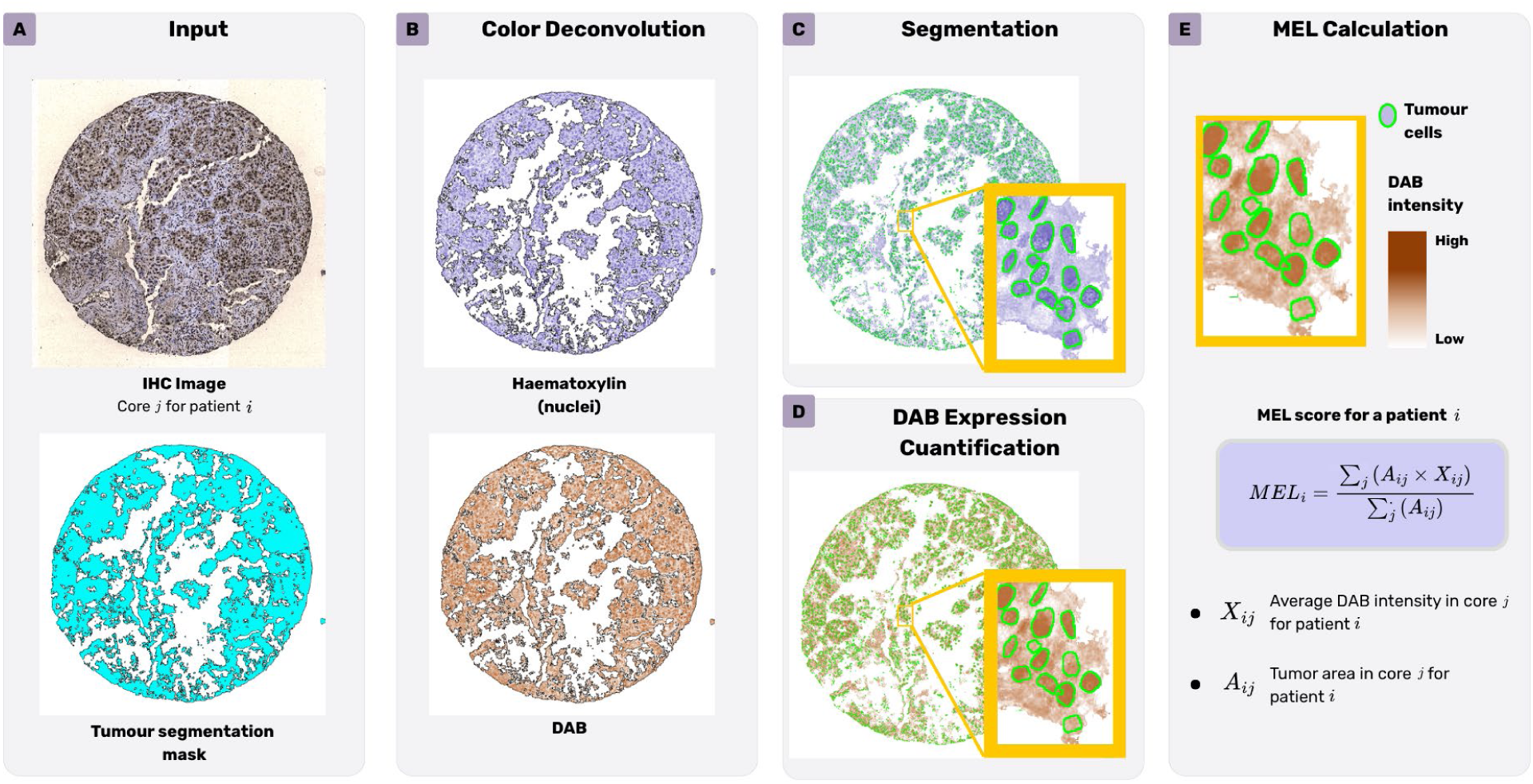
Outline of the marker quantification process for MEL calculation. BRCA1 marker was used in this example.

Cytoplasmic QKI was quantified using the same procedure, after excluding the nuclear regions from the final region of interest. For membrane GLUT1, tumour nuclei were first segmented as described above and used as seeds for a marker-controlled watershed algorithm applied to the inverted DAB channel. The resulting watershed lines were slightly dilated to define the membrane regions, from which mean DAB intensity was measured. The patient-level MEL score was calculated using the mean membrane staining intensity and tumour area of each available core.

### Open-Source Quantification Toolbox

An open-source Fiji plugin was developed to automate the quantification of nuclear, cytoplasmic, and membrane-associated IHC markers. The toolbox supports both single-image and batch processing and allows users to select the cellular compartment to be analysed and the region of interest. The region of interest can comprise the whole tissue, a tissue region selected from an input segmentation mask, or a manually defined region for single-image analysis.

For each image, the plugin displays the analysed tissue region and reports its area, mean DAB intensity in the selected cellular compartment, standard deviation, mode, minimum, and maximum values. In batch mode, the results are automatically exported to an Excel or CSV file. Further details of the plugin usability can be found in Supplementary Material.

The MEL quantification plugin is available at https://github.com/UPNa-Biomedical-Engineering-Group/MEL-quantification.

### TMA Batch Normalization

A TMA-based normalization procedure was applied to reduce technical variation in staining intensity between TMAs, ensuring that the differences observed were truly due to biological or physiological processes [25], [26]. Inter-TMA variability was assessed using the coefficient of variation (CV), calculated as the ratio between the standard deviation and mean staining intensity. According to the criteria adopted from [27], normalization was considered unnecessary for CV values below 20%, optional for values between 20% and 30%, and strongly recommended for values above 30%.

For markers requiring normalization, a scaling factor was applied to the MEL score:

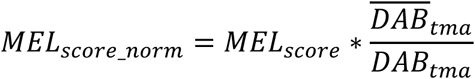

where DAB_tma_ is the mean DAB intensity of the TMA from which the image originated, and DAB_tma_ is the mean of the six TMA-specific DAB values.

### Prognostic Signature

Survival was assessed over a 60-month follow-up period using disease-free survival (DFS), defined as the time from surgery to cancer relapse, and overall survival (OS), defined as the time from surgery to death. As in our previous publication [6], a combined prognostic index (CPI) incorporating biomarker expression and pathological tumour stage was calculated.

First, a MEL-based prognostic index (PI) was obtained using Cox regression models including the MEL scores of the three markers for each survival endpoint:

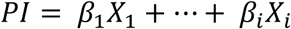

where the *β_i_* represents the strength and the direction of the association between the i-th covariate and the survival outcome, and X_i_ is the corresponding patient-specific value.

The CPI was then calculated using a bivariate Cox regression model including the PI and tumour stage:

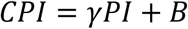

where *γ* is the scaling coefficient adjusting the contribution of the PI to the CPI, and *B* is the stage- specific coefficient.

Prognostic performance was evaluated using Harrell’s concordance index (C-index) and Kaplan– Meier survival curves, with the CPI dichotomized at the median and groups compared using the log- rank test.

## RESULTS

### Segmentation accuracy

Segmentation performance was primarily evaluated using the Dice coefficient, complemented by precision, recall, volumetric similarity, and object-level analyses (Table 2). The model achieved excellent agreement for the background region (Dice = 0.986 ± 0.013, precision = 0.987 ± 0.011, recall = 0.985 ± 0.019) and robust and comparable performance for the two major tissue regions, tumour (Dice = 0.694 ± 0.169, precision = 0.697 ± 0.171, recall = 0.743 ± 0.212) and stroma (Dice = 0.698 ± 0.166, precision = 0.726 ± 0.165, recall = 0.720 ± 0.213), with high volumetric similarity (0.848 ± 0.143 and 0.852 ± 0.147, respectively) and object detection rates of 58.1% and 64.3%.

**Table 2:** Evaluation of tissue compartment segmentation. Data are presented as mean ± standard deviation across all evaluated images, except for Object Detection Rate (Obj. DR), which represents the proportion of target connected components correctly identified using an overlap threshold of ≥50% against ground truth. Abbreviations: GT, ground truth; Pred., predicted; Vol. Sim., volumetric similarity; Obj. DR, object detection rate.

| **Tissue Compartment** | **Dice** | **Precision** | **Recall** | **Vol. Sim.** | **Obj. DR (%)** | **GT Area (%)** | **Pred. Area (%)** |
|---|---|---|---|---|---|---|---|
| Background | 0.986 ± 0.013 | 0.987 ± 0.011 | 0.985 ± 0.019 | 0.994 ± 0.008 | 55.8 | 40.28 ± 4.71 | 40.13 ± 4.24 |
| Tumour | 0.694 ± 0.169 | 0.697 ± 0.171 | 0.743 ± 0.212 | 0.848 ± 0.143 | 58.1 | 27.40 ± 10.46 | 29.61 ± 13.31 |
| Stroma | 0.698 ± 0.166 | 0.726 ± 0.165 | 0.720 ± 0.213 | 0.852 ± 0.147 | 64.3 | 28.48 ± 9.69 | 28.78 ± 11.87 |
| Immune Infiltrate | 0.171 ± 0.251 | 0.432 ± 0.406 | 0.191 ± 0.250 | 0.262 ± 0.330 | 37.2 | 3.83 ± 5.26 | 1.48 ± 2.97 |

The immune infiltration class achieved a lower Dice coefficient (0.171 ± 0.251), reflecting its intrinsic complexity. This class represented only 3.83% of the annotated tissue area and comprised numerous very small or highly fragmented regions. Object-level analysis showed that although only 37.2% of annotated objects reached the 50% overlap criterion, performance was strongly size-dependent: intermediate-sized objects (11–5000 pixels), representing over half of all annotations, achieved a detection rate of 61.2% with a mean overlap of 61.4%, whereas extremely small (≤10 pixels) and very large (≥5000 pixels) objects were substantially more challenging. These findings indicate that the low Dice score is primarily driven by severe class imbalance and the heterogeneous morphology of immune infiltrates rather than by a failure to localize biologically relevant immune regions.

Overall, the segmentation framework demonstrated robust performance in distinguishing key histological compartments, particularly tumour regions, which are central to downstream MEL score computation.

### TMAs batch normalization

A difference in the average staining intensities was observed between the first three TMAs (1,2,3) and the last three ones (4,5,6) as they were prepared and immunostained in separate batches. Hence, this technical difference across TMAs is a key factor to take into account in the quantification phase. This is shown in Figure 3A, where there is an offset between MEL and visual H-score of GLUT1 in TMAs 1-2-3 that is not observed in TMAs 4-5-6. Since MEL is an objective measure, it provides evidence of a genuine staining variation between the two batches of TMAs. Consequently, the absence of a discrepancy between average visual H-scores of the initial and final TMAs suggests that the observers adjusted their evaluation criteria for each batch artificially, thereby accounting for the existing, undesired staining difference. This effect is corrected after applying TMA normalization to GLUT1 marker, as shown in Figure 3B.

**Figure 3:**
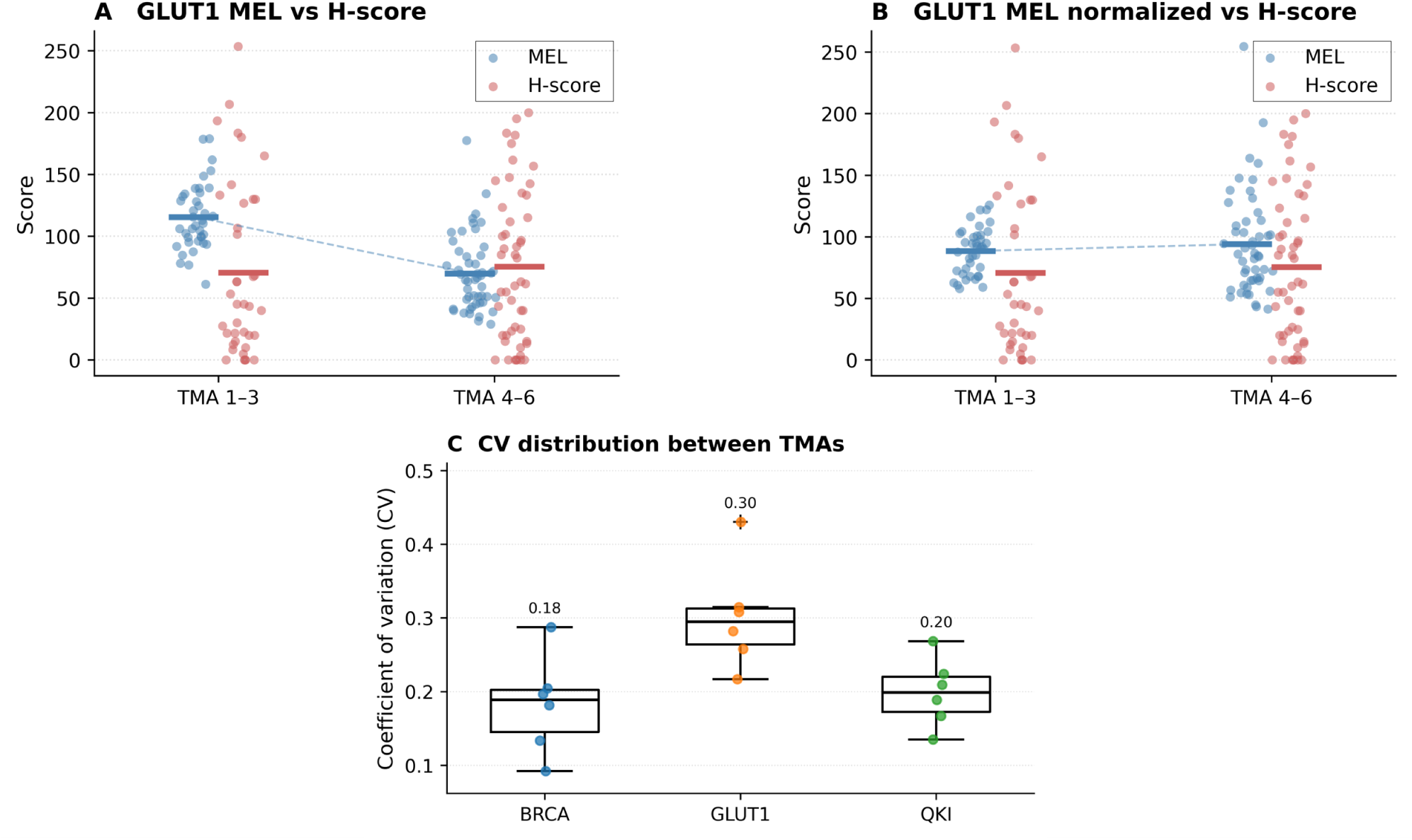
Comparison between the first batch (TMAs 1,2 and 3) and the second batch (TMAs 4,5 and 6) for GLUT1 marker. A) H-score vs. not normalized MEL score. B) H-score vs. MEL score after normalization. Individual points correspond to individual cores, solid line indicates mean of the distribution. C) Distribution analysis of the coefficient of variance (CV) of DAB staining across TMAs for the three markers.

The CVs of each of the 6 TMAs for each marker are shown in Figure 3C. CV values ranged from 0.10 to 0.45, indicating varying levels of staining variability across TMAs. Specifically, mean CV was 0.18 for BRCA1, indicating consistent staining across TMAs, 0.30 for GLUT1, indicating substantial variability in staining across TMAs, and 0.20 for QKI, indicating low/moderate variability. Based on these results, our choice was to apply normalization only to GLUT1.

### Quantification of marker expression

To validate the proposed automatic image processing pipeline, a statistical correlation analysis was carried out between visual H-Scores of all patients, manually assessed by experts in Martinez-Terroba et al. [6], and our quantified MEL scores. This correlation analysis quantifies the relationship between two variables in terms of strength and direction. Specifically, the Spearman’s correlation coefficient was used to assess the strength of the association between both scores. All analyses yielded statistically significant correlations between MEL scores and visual H-scores, thus validating the use of the MEL score as an automatic alternative to the well-established H-score (Figure 4). Indeed, statistically significant correlation was found for BRCA1 in tumoral cell nuclei (rho = 0.690, p < 0.001), for QKI in tumoral cell nuclei and cytoplasm (rho = 0.623, p < 0.001, and rho = 0.612, p < 0.001, respectively), and for GLUT1 in cell membrane (rho = 0.623, p < 0.001). Remarkably, GLUT1 staining normalization across the six TMAs improved the correlation of the MEL score with the visual H-score from a rho of 0.391 to 0.623.

**Figure 4:**
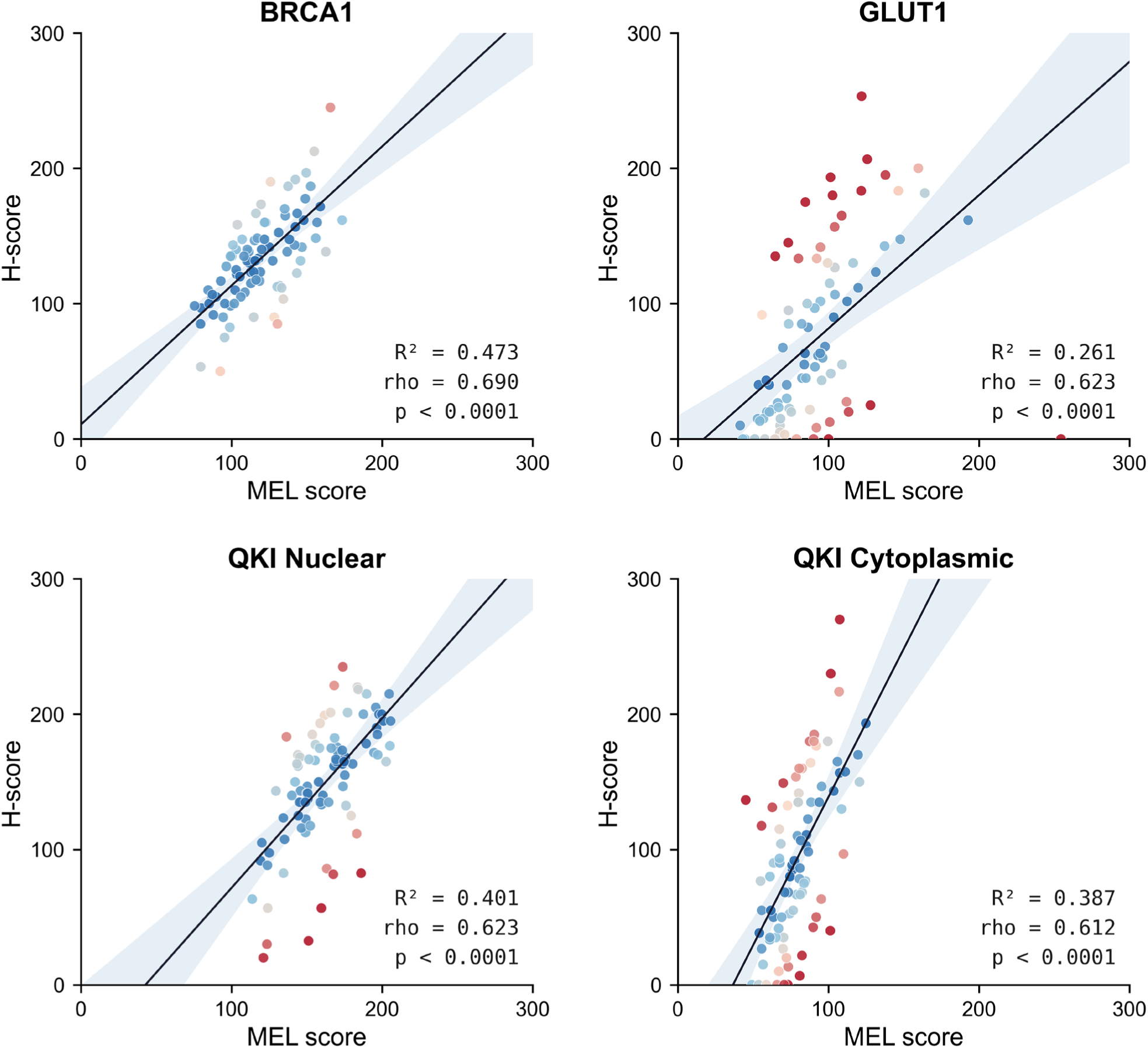
Correlation analysis of each prognostic marker between automatic MEL score and visual H-score. Spearman’s correlation coefficient was used to assess the strength of the association between both scores.

### Replication and validation of prognostic signature

Once the MEL values of each marker were obtained, the CPI was obtained for each patient by means of a bivariate cox regression. Given the distinct biological and clinical implications of tumour recurrence (DFS) and patient mortality (OS), two separate bivariate Cox proportional hazard regression models were fitted:

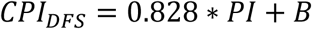

where B is a coefficient that changes for each tumour stage (IA, B=0; IB, B=1.091; IIA, B=1.391; IIB, B=1.923; and IIIA, B=1.177).

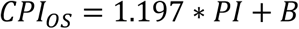

where B changes for each tumour stage (IA, B=0; IB, B=1.091; IIA, B=0.997; IIB, B=2.723; and IIIA, B=0.825).

The survival curves comparing the visual and automatic prognostic models can be seen in Figure 5. The Kaplan-Meier survival analysis for DFS showed a statistically significant difference between the two prognostic patient groups defined both by the visual H-score and the automatic MEL score (Figure 5 top row). Patients classified in the high combined prognostic index (HCPI) group had a significantly lower DFS compared to those in the low combined prognostic index (LCPI) group. The log-rank test confirmed the significance of this difference both for the CPI based on automatic marker scoring (p < 0.001) and the CPI based on visual marker scoring (p < 0.001). Moreover, the hazard ratio (HR) estimated with the automatic prognostic model was 4.43 (95% CI: 1.96 – 10.02), indicating that patients in the HCPI group had an approximately fourfold increased risk of disease recurrence or progression compared to those in the LCPI group. A similar result was obtained with the manual - visual- prognostic model (HR = 4.24; 95% CI: 1.88 – 9.54). These results suggest that the combined prognostic model for DFS built from an automatic MEL score is effective, allowing the stratification of patients based on their likelihood of experiencing disease recurrence. Moreover, patients classified in the HCPI group had a significantly lower OS compared to those in the LCPI group both for the visual CPI (p < 0.01) and the automatic CPI (p < 0.05) (Figure 5 bottom row). The estimated hazard ratios, HR = 6.35 (95% CI: 1.91 – 21.08) for the visual CPI and HR = 5.12 (95% CI: 1.57 – 16.73) for the automatic CPI, suggest that patients with higher CPI scores are at a significantly increased risk of 5 year-mortality compared to those with lower scores (5 to 6 times higher).

**Figure 5:**
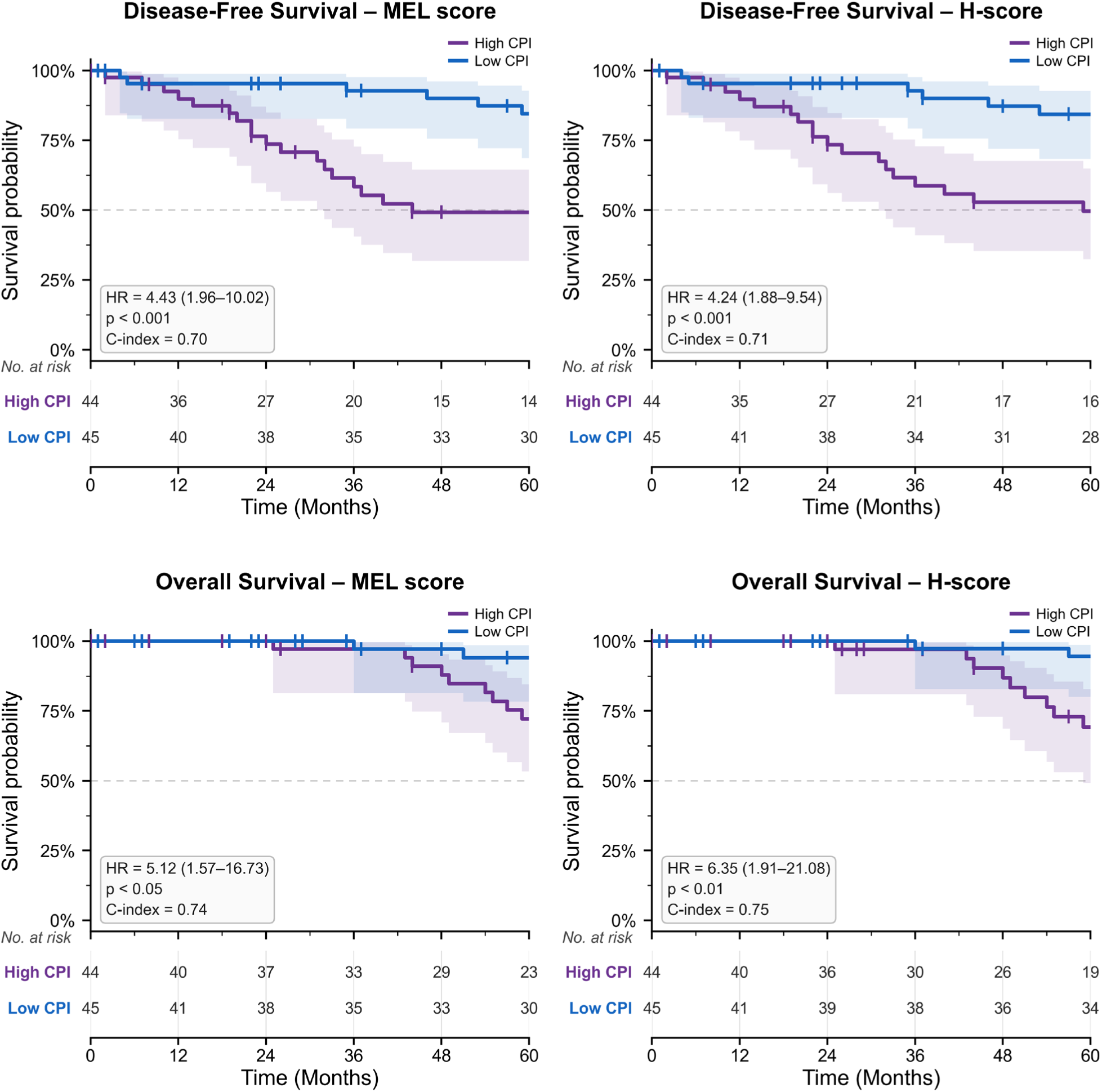
Survival Analysis for DFS and OS events: on the left the results obtained with a prognostic model using the automatic MEL scores and on the right the results obtained with a prognostic model using the visual H-scores.

The C-index was used to evaluate the ability of the models to correctly rank patients according to their recurrence or survival probabilities. The obtained C-index values indicated moderate to good discriminatory ability for all the models, with values ranging from 0.70 to 0.75. Specifically, the prognostic models for DFS achieved C-indexes of 0.70 and 0.71 using the automatic MEL score and the visual H-score, respectively, whereas the prognostic models for OS achieved C-indexes of 0.74 and 0.75 using the automatic MEL score and visual H-score, respectively. These results suggest that the automatic pipeline performs similar to the visual assessment in the construction of prognostic models for distinguishing patients with different recurrence or survival probabilities. While the

Kaplan-Meier analysis demonstrated a favourable outcome by effectively stratifying patients into distinct risk groups, the C-index values reinforce the models’ ability to rank individuals based on their recurrence or survival risk with a reasonable degree of accuracy.

## DISCUSSION

An automatic metric to quantify marker expression in IHC images is essential for enhancing the objectivity, reproducibility, and scalability of histopathological analysis. Traditional visual assessment methods, such as the visual H-score, rely heavily on the subjective interpretation of pathologists, introducing variability that may affect the consistency and reliability of the results, particularly when processing large datasets or conducting multi-center studies. In contrast, the automatic metric proposed in this study, the MEL score, offers a robust alternative by providing a continuous, standardized measure of staining intensity within tumoral regions. The MEL score circumvents the limitations of manual grading by leveraging image analysis algorithms to extract quantitative features directly from digitized tissue slides. This eliminates inter-observer variability and enables high-throughput processing, which is particularly valuable in modern digital pathology workflows. Furthermore, the MEL score demonstrated a strong and statistically significant correlation with the visual H-score across all considered markers, indicating its validity as a surrogate metric.

Beyond reproducing the H-score, the MEL framework introduces several methodological improvements. First, MEL incorporates automated segmentation of distinct tissue regions, enabling tumour-specific quantification while excluding non-tumour tissue from the assessment. The segmentation showed consistent performance for the major tissue compartments, particularly tumour and stroma, whereas the lower performance for immune infiltration reflected its sparse, heterogeneous, and fragmented morphology. Importantly, object-level analysis indicated better performance for immune infiltrates of intermediate size, suggesting that the overall Dice score does not fully capture the model’s ability to identify relevant immune regions. Building on this tissue- aware segmentation, MEL incorporates tumour-area weighting across multiple tissue cores, providing a more representative estimate of biomarker expression within the sampled tissue. Although TMAs are primarily research tools rather than part of routine pathological practice, this approach addresses a broader challenge that is also relevant to clinical assessment: intratumour heterogeneity in biomarker expression across different tumour regions, blocks, or tissue sections from the same patient. By enabling objective quantification across multiple samples, MEL could facilitate the assessment of such heterogeneity and help distinguish regional differences in biomarker expression from variability introduced by subjective scoring. Second, the proposed TMA batch normalization strategy effectively compensated for inter-TMA staining differences, particularly for GLUT1, demonstrating the ability of our method to address technical variability in retrospective research cohorts. An additional strength of the framework is its flexibility across cellular compartments. While many automated approaches focus primarily on nuclear biomarkers, MEL successfully quantified nuclear, cytoplasmic, and membrane-associated markers. In particular, the dedicated segmentation strategy developed for GLUT1 membrane staining improved agreement with visual assessment after normalization. Given the biological relevance of GLUT1 as a surrogate marker of tumour metabolic activity, this approach may also be applicable to other tumour types in which metabolic reprogramming plays a key prognostic role. Importantly, MEL was not only validated against visual scoring but also reproduced a previously established and clinically validated prognostic signature with comparable discriminatory performance. The resulting prognostic models achieved similar hazard ratios, survival stratification, and concordance indices to those obtained using expert-derived H-scores. These findings suggest that MEL preserves the biological and clinical information captured by conventional assessment while providing a more objective and reproducible measurement framework.

The implementation of the methodology as an open-source, accessible Fiji plugin additionally facilitates its adoption by the research community and supports its potential translation into routine digital pathology workflows outside the research-intended TMA setting. By integrating the MEL score into these workflows, pathologists can benefit from a more consistent and efficient evaluation of biomarker expression. This automation not only supports clinical decision-making but also lays the foundation for large-scale biomarker studies and the development of AI-assisted diagnostic tools. Future studies should evaluate the performance of MEL in larger independent cohorts, with alternative and even more accurate prognostic factors and across different institutions to further assess its generalizability and clinical robustness. Overall, the MEL score represents a significant step toward the standardization of IHC quantification, fostering greater confidence in the prognostic and predictive value of tissue-based biomarkers.

## CODE AVAILABILITY

The code for the whole MEL calculation pipeline is publicly available. The TSS tissue segmentation plugin can be found in https://github.com/CVPD/Trainable_Superpixel_Segmentation. The MEL quantification pipeline is available at https://github.com/UPNa-Biomedical-Engineering-Group/MEL-quantification.

## Data Availability

The code for the whole MEL calculation pipeline is publicly available. The TSS tissue segmentation plugin can be found in https://github.com/CVPD/Trainable_Superpixel_Segmentation. The MEL quantification pipeline is available at https://github.com/UPNa-Biomedical-Engineering-Group/MEL-quantification.
The images are available upon reasonable request to the corresponding authors.

## ACKNOWLEDGEMENTS

The authors would like to thank all the subjects who participated in this study. MA was supported by the Public University of Navarre (grant number PJUPNA2026-12555). COS was supported by the Ministry of Science, Innovation and Universities of Spain (MICIU/AEI/10.13039/501100011033) and by the European Union through the FEDER (a way to make Europe) program under projects PID2024-155384OB-C22 and PID2021-122409OB-C22. LMM was supported by the Instituto de Salud Carlos III (ISCIII) through the Strategic Action in Health (AES 2025) call, under grant number PI25/00123, and co-funded by the European Union. He also receives funding from the European Union’s Horizon Europe Mission Cancer Programme under GA101136552 SPACETIME. AC was supported by Ministerio de Ciencia, Innovación y Universidades (PID2025-175425OB-I00) and Worldwide Cancer Research Foundation. This work was partially supported by grants PID2024- 157485NB-I00, RED2024-153844-T and AIA2025-164165-C42 (to I.A-C.) funded by MICIU/AEI/10.13039 /501100011033 and by FEDER, UE. We gratefully acknowledge financial support (to I.A-C.) from Gobierno Vasco-Eusko Jaurlaritza (IT2001-26). MS received a PhD grant from the Program of Grants attached to Projects of The University of Navarra. Views and opinions expressed are however those of the author(s) only and do not necessarily reflect those of the European Union or HADEA. Neither the European Union nor the granting authority can be held responsible for study design, data collection, data analysis, data interpretation, or writing of the report.

## AUTHORS CONTRIBUTION STATEMENT

FC, MS, AI, JS, IAC and MA contributed to software development; FC, MS, AI, IAC, COS and MA contributed to data analysis; FC, MS, DO, IAC, AC, LMM, COS and MA contributed to data interpretation; DO, CS and AC contributed to image annotation; DO, CS, AC and LMM contributed to image and data curation; IAC, AC, LMM, COS and MA contributed to project supervision and funding acquisition; all authors contributed to the writing of the manuscript.

## DATA AVAILABILITY STATEMENT

The images are available upon reasonable request to the corresponding authors.

## ETHICS APPROVAL STATEMENT

The project was approved by the University of Navarra Ethics Committee (Ref. 092.2012 mod 1).

## PATIENT CONSENT STATEMENT

Patients signed the informed consent required by the Ethics Committee.

