## Supplementary Material for "Development and Validation of an Automated Image-Based Method for Prognostic Stratification in Lung Adenocarcinoma using Immunohistochemical Markers"

### SUPPLEMENTARY MATERIALS AND METHODS

#### Patient Cohort and Clinicopathological Assessment

Lung adenocarcinoma tumour samples were obtained from a retrospective cohort of patients treated at Clínica Universidad de Navarra (CUN, Pamplona, Spain) between 2002 and 2024. The cohort comprised 128 patients with surgically resected stage I–III non-small cell lung cancer, including current smokers, former smokers, and never smokers. Patients were eligible if they underwent surgical resection at CUN and had no history of cancer within the 5 years preceding surgery, except for non-melanoma skin cancer. The study was conducted under an approved ethical protocol (ref. 092.2012 mod 1; approved 06/09/2022).

Formalin-fixed paraffin-embedded tissue blocks were used to construct six tissue microarrays (TMAs). Three consecutive 4-µm sections were obtained from the TMAs and immunostained for BRCA1, GLUT1, and QKI, the three biomarkers comprising the previously reported prognostic signature [6]. Three tumour cores per patient were analysed for each marker. IHC images were acquired in RGB format using a Leica Aperio image scanner with a 20× objective.

In the original study, IHC expression was quantified using visual H-scores, which were integrated into a multivariable Cox regression-derived prognostic index [6]. For the present study, patients were eligible only when at least one TMA core image was available for each biomarker and visual H-scores were available for all three markers. Of the initial 128 patients, 96 fulfilled these requirements and constituted the final analytical cohort. Detailed information on histopathological classification of lung adenocarcinoma subtypes, selection of representative tumour regions, TMA construction, immunohistochemistry, including antigen retrieval and antibody validation, and visual H-score assessment has been previously reported [6].

Disease-free survival was defined as the time from surgery to the first documented recurrence or death from any cause. Overall survival was defined as the time from surgery to death from any cause. Patients without an event were censored at the date of the last available follow-up.

**Trainable Superpixel Segmentation for Tissue Region Delineation**

To enable automated analysis of histological regions of interest, tissue compartment segmentation was performed using the Trainable Superpixel Segmentation (TSS) Fiji/ImageJ plugin developed by the Computer Vision and Pattern Discovery (CVPD) group [19]. The plugin implements a supervised image segmentation framework based on the classification of image superpixels rather than individual pixels, thereby reducing computational complexity while preserving tissue-level contextual information. The software is distributed as an open-source Fiji plugin and integrates functionality from ImageJ/Fiji, Weka, and MorphoLibJ.

**Superpixel-Based Segmentation Strategy**

The TSS workflow consists of four main stages: (i) image over-segmentation into superpixels, (ii) extraction of descriptive image features for each superpixel, (iii) supervised classifier training from annotated examples, and (iv) prediction of tissue classes in previously unseen images. The methodological framework is described in detail in the original technical report accompanying the software. Superpixels are defined as spatially coherent groups of neighboring pixels with similar visual characteristics, allowing biologically meaningful structures to be represented by compact image primitives rather than isolated pixels. Please note that the calculation of superpixels is external to the plugin and can be done using any other available tool/plugin such as SLIC or watershed.

**Feature Extraction**

The plugin extracts intensity-based statistical features from each superpixel using the Intensity Measurements functionality of MorphoLibJ. These include measurements such as mean, median, standard deviation, minimum, maximum and kurtosis, among other statistical descriptors. The same measurements are also computed for neighbouring (touching) superpixels, providing local contextual information. The resulting feature vectors are used to train supervised Weka classifiers and assign class labels to superpixels.

**Classifier Training and Prediction**

TSS adopts a supervised learning approach in which training examples are provided from annotated image data and used to train a classifier implemented through the Weka framework. Once trained, the resulting model assigns a tissue class label to each superpixel in new images acquired under comparable staining and imaging conditions. The framework supports multiclass segmentation and can therefore distinguish several histological compartments simultaneously.

**Relevance for Histopathological Image Analysis**

The superpixel-based approach is particularly suitable for histopathology because it enables classification at the level of small tissue regions rather than individual pixels. In immunohistochemical tissue microarrays, tumor regions frequently contain heterogeneous mixtures of malignant cells, inflammatory infiltrates, stromal tissue, and background structures. By incorporating statistical information from both superpixels and their local neighborhoods, TSS can capture contextual information that contributes to the discrimination of histological tissue compartments.

For the analyses described in this study, the resulting segmentation masks were used to restrict biomarker quantification to tumor regions of interest, thereby excluding non-tumor tissue from downstream measurements and improving the specificity of the derived immunohistochemical metrics.

**Open-Source MEL Quantification Toolbox**

An open-source Fiji macro toolbox was developed on top of the TSS plugin to automate the quantification of the nuclear, cytoplasmic and membrane IHC markers described in the main text. This toolbox could be used in clinical practice to streamline the automation of these or other markers with cellular compartment information (nucleus, cytoplasm or membrane). The plugin allows choosing different modes of operation:

- **Single image mode:** one image at a time is analysed.
- **Batch mode:** an entire folder of images is analysed.

Each of these modalities has an additional set of options that the user can specify. These include the choice of the cell compartment (nuclei, membrane or cytoplasm) to be analysed and how the region of interest will be selected:

- **Whole tissue:** the entire tissue is analysed, without distinguishing between different tissue regions (e.g., healthy and stromal tissue).
- **ROI from mask:** an input segmentation mask, required also as input, is used to select and analyse only the cells in the tissue region of interest (e.g., tumoral tissue).

Furthermore, for single image mode only, there is an additional option that can be selected:

- **Manual ROI:** a manual ROI can be drawn to analyse only the cells located in the selected tissue region. The user can manually draw one or several, disjoint ROIs.

For each modality, the original image is shown with an overlay of the tissue regions being analysed, and a Fiji window with the quantified results is displayed, namely the area of the selected tissue region, the average DAB intensity in the selected cellular compartments of the ROI, as well as complimentary metrics as the standard deviation, mode, min and max values of DAB in that compartment. In batch mode, all these metrics are calculated and saved in an output excel or csv file.

**Software Availability**

Both the Trainable Superpixel Segmentation plugin and the MEL-quantification toolbox are freely available through the CVPD[1] and the UPNa Biomedical Engineering Group[2] repositories and can be executed within Fiji/ImageJ[3]. The software relies on open-source libraries for image handling (ImageJ/Fiji), machine learning (Weka[4]) and feature extraction (MorphoLibJ[5]).

[1] https://github.com/CVPD/Trainable_Superpixel_Segmentation
[2] https://github.com/UPNa-Biomedical-Engineering-Group/MEL-quantification
[3] https://fiji.sc
[4] http://www.cs.waikato.ac.nz/ml/weka/
[5] https://imagej.net/plugins/morpholibj
